# Impact of Pre-Existing Chronic Viral Infection and Reactivation on the Development of Long COVID

**DOI:** 10.1101/2022.06.21.22276660

**Authors:** Michael J. Peluso, Tyler-Marie Deveau, Sadie E. Munter, Dylan Ryder, Amanda Buck, Gabriele Beck-Engeser, Fay Chan, Scott Lu, Sarah A. Goldberg, Rebecca Hoh, Viva Tai, Leonel Torres, Nikita S. Iyer, Monika Deswal, Lynn H. Ngo, Melissa Buitrago, Antonio Rodriguez, Jessica Y. Chen, Brandon C. Yee, Ahmed Chenna, John W. Winslow, Christos J. Petropoulos, Amelia N. Deitchman, Joanna Hellmuth, Matthew A. Spinelli, Matthew S. Durstenfeld, Priscilla Y. Hsue, J. Daniel Kelly, Jeffrey N. Martin, Steven G. Deeks, Peter W. Hunt, Timothy J. Henrich

**Affiliations:** Division of HIV, Infectious Diseases, and Global Medicine, University of California, San Francisco, CA, USA; Division of Experimental Medicine, University of California, San Francisco, CA, USA; Department of Epidemiology and Biostatistics, University of California, San Francisco, CA, USA; Monogram Biosciences Inc., South San Francisco, CA, USA; School of Pharmacy, University of California, San Francisco CA, USA; Department of Neurology, University of California, San Francisco, CA, USA; Division of Cardiology, University of California, San Francisco, CA, USA

**Keywords:** COVID-19, SARS-CoV-2, post-acute sequelae of SARS-CoV-2 infection (PASC), Long COVID, Epstein-Barr Virus, Cytomegalovirus, HIV

## Abstract

The presence and reactivation of chronic viral infections such as Epstein-Barr virus (EBV), cytomegalovirus (CMV) and human immunodeficiency virus (HIV) have been proposed as potential contributors to Long COVID (LC), but studies in well-characterized post-acute cohorts of individuals with COVID-19 over a longer time course consistent with current case definitions of LC are limited. In a cohort of 280 adults with prior SARS-CoV-2 infection, we observed that LC symptoms such as fatigue and neurocognitive dysfunction at a median of 4 months following initial diagnosis were independently associated with serological evidence of recent EBV reactivation (early antigen-D [EA-D] IgG positivity) or high nuclear antigen IgG levels, but not with ongoing EBV viremia. Evidence of EBV reactivation (EA-D IgG) was most strongly associated with fatigue (OR 2.12). Underlying HIV infection was also independently associated with neurocognitive LC (OR 2.5). Interestingly, participants who had serologic evidence of prior CMV infection were less likely to develop neurocognitive LC (OR 0.52) and tended to have less severe (>5 symptoms reported) LC (OR 0.44). Overall, these findings suggest differential effects of chronic viral co-infections on the likelihood of developing LC and predicted distinct syndromic patterns. Further assessment during the acute phase of COVID-19 is warranted.

**SUMMARY:** The authors found that Long COVID symptoms in a post-acute cohort were associated with serological evidence of recent EBV reactivation and pre-existing HIV infection when adjusted for participant factors, sample timing, comorbid conditions and prior hospitalization, whereas underlying CMV infection was associated with a decreased risk of Long COVID.

## BACKGROUND

Intense efforts are underway to understand the pathophysiologic mechanisms that drive Long COVID (LC), a type of post-acute sequelae of SARS-CoV-2 infection (PASC) characterized by persistent or recurrent symptoms that interfere with quality of life (1, 2). Prior work has identified immune activation (3, 4), microvascular dysfunction (5, 6), autoimmunity (7, 8), and SARS-CoV-2 viral persistence (9–12) as mechanisms potentially contributing to LC. However, not all studies have confirmed these processes (13, 14), and identification of the determinants of PASC is essential to efforts to prevent and treat this condition (15).

Latent Epstein-Barr virus (EBV) is a ubiquitous human herpesvirus harbored by the vast majority (90-95%) of adults in high-income settings, usually defined by the presence of detectable EBV viral capsid antigen (VCA) IgG levels (16). EBV can reactivate in immunocompromised individuals, as well as in the setting of physiologic stressors including acute infection (17). In some cases, EBV reactivated in tissues may not manifest with detectable circulating DNA in blood (18, 19). While reactivation of EBV is often considered to be a marker of physiologic stress rather than an independent pathophysiologic process, recent studies have demonstrated that EBV infection may drive multiple sclerosis (20), perhaps due to aberrant autoreactive immune responses to viral infection (21). Prior studies have demonstrated EBV reactivation, as defined by detectable circulating EBV DNA or EBV VCA IgM positivity, during acute SARS-CoV-2 infection (22–26). However, these studies typically involved hospitalized patients and the high rates of reactivation (e.g., >80% of patients) were observed primarily in those receiving positive-pressure ventilation or other ICU-level care. Furthermore, VCA IgM levels wane rapidly and may not be useful outside the context of acute or subacute SARS-CoV-2 infection.

EBV reactivation has also been proposed as a driver of Long COVID. One small but highly provocative study identified EBV early antigen-diffuse (EA-D) IgG positivity, a marker of recent viral activity or reactivation, among two-thirds of individuals experiencing LC (27). EBV EA-D IgG levels were higher in those with more PASC symptoms. EBV EA-D IgG levels rise early after recent viral activity like VCA IgM levels but remain positive for months prior to decaying to undetectable levels (VCA IgG levels remain elevated indefinitely) (28). As a result, EBV EA-D IgG levels may act as a surrogate for recent EBV reactivation in tissues several months after the reactivation event (**Figure 1**). EBV also elicits life-long nuclear antigen (NA) IgG responses, which initially increase at the time of transition between the lytic and latent phases of acute EBV infection (28). Given a several-month lag in NA IgG responses following viral activity, it is possible that increases in NA IgG levels sampled months following COVID-19 onset in convalescent LC cohorts may act as a potential marker of EBV reactivation or other inflammatory insult at the time of acute SARS-CoV-2 infection. More recent work has shown that EBV DNA detectability during acute SARS-CoV-2 infection predicted the presence of symptoms at 30-60 days post-COVID (7). Although limited by small sample size, sex imbalance, and over-representation of hospitalized individuals, as well as relatively short duration of follow-up, these studies suggest that further investigation of the relationship between EBV-related pathology and Long COVID is warranted. Also needed are studies controlling for potentially confounding factors in the interpretation of EBV reactivation and underlying chronic viral infections, such as timing of sample collection, hospitalization and severity of disease during initial infection, underlying health conditions, and other participant demographics, as well as studies accounting for the heterogeneity in syndromic patterns of LC that may reflect different disease phenotypes potentially caused by pathophysiologic mechanisms.

**Figure 1.**
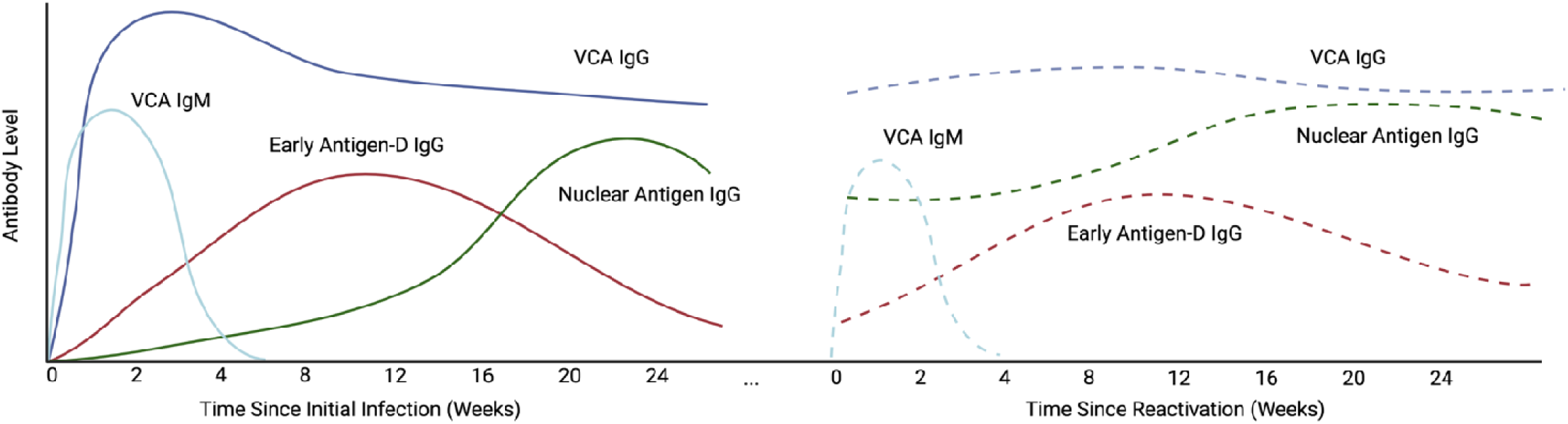
Schema of EBV-specific antibody responses during acute infection and hypothetical responses during SARS-CoV-2-related reactivation. EBV viral capsid antigen (VCA) IgM and IgG rise fairly early after acute infection, with VCA IgG levels persisting long-term. EBV nuclear antigen (NA) IgM levels rise more slowly following acute infection, at a time when virus changes from the lytic to latent phase of infection. Early antigen-D (EA-D) IgG responses rise early following acute infection but decay, often to low or undetectable levels over many months. The dashed lines represent potential changes to antibody levels following EBV reactivation secondary to an insult such as acute SARS-CoV-2 infection. EBV EA-D IgG responses and perhaps increases in baseline levels of EBV NA IgG may be observed 3-4 months following reactivation.

Given the potential connection between EBV reactivation and the development of Long COVID, there is also now much interest in how other underlying chronic viral infections, such as cytomegalovirus (CMV) and human immunodeficiency virus (HIV), may influence both acute SARS-CoV-2 infection and post-acute sequelae. For example, CMV seropositivity may be associated with more severe acute initial infection (29, 30), but it is not known whether CMV plays a significant role in Long COVID. Recent data also demonstrated a potential link between the development of T cell receptor sequence repertoires suggesting CMV cytolytic activity associated with gastrointestinal symptoms up to 2 months following acute infection (7), but direct evidence of CMV infection and LC are lacking. Similarly, we and others have recently observed that people with HIV may have a greater risk of developing LC (31, 32), but larger studies that control for factors such as human herpesvirus infections (many of which are enriched in people with HIV), participant demographics, and other underlying health conditions in both hospitalized and non-hospitalized participants are urgently needed.

In this study, we sought to investigate the prevalence of underlying CMV and HIV infection and evidence of EBV reactivation in a well-characterized post-acute COVID-19 cohort of individuals with and without various Long COVID symptoms (e.g. fatigue, neurocognitive, cardiopulmonary, gastrointestinal) approximately four months following initial SARS-CoV-2 infection. We evaluated the independent associations between pre-existing CMV and EBV reactivation and a variety of different LC symptom groups controlled for clinical and demographic factors, including underlying HIV infection and details about acute infection. We hypothesized that the group experiencing LC symptoms would be enriched for evidence of EBV reactivation and underlying CMV seropositivity in comparison to individuals reporting complete recovery from COVID-19.

## RESULTS

### Relationship between participant factors and Long COVID symptoms

Participant demographics, pre-existing health conditions, COVID-19-related hospitalization and EBV antibody test results were compared by LC symptom group in 280 participants at the time point beyond 60 days that was closest to 4 months (median 123 days) following nucleic acid-based diagnosis of acute SARS-CoV-2 infection with available data as shown in **Table 1**. Overall, the median age was 45 years, 56% were men at birth, 18% had been hospitalized during acute infection, 65% had a body mass index (BMI) of >30, and 19% were living with HIV (the cohort was deliberately enriched for such individuals). In univariate analyses, there were significantly higher proportions of participants with LC or severe LC (reporting more than 5 symptoms, LC>5) who had been hospitalized compared to those without LC (21% and 26% versus 9%, respectively; all P<0.05).

**Table 1.**
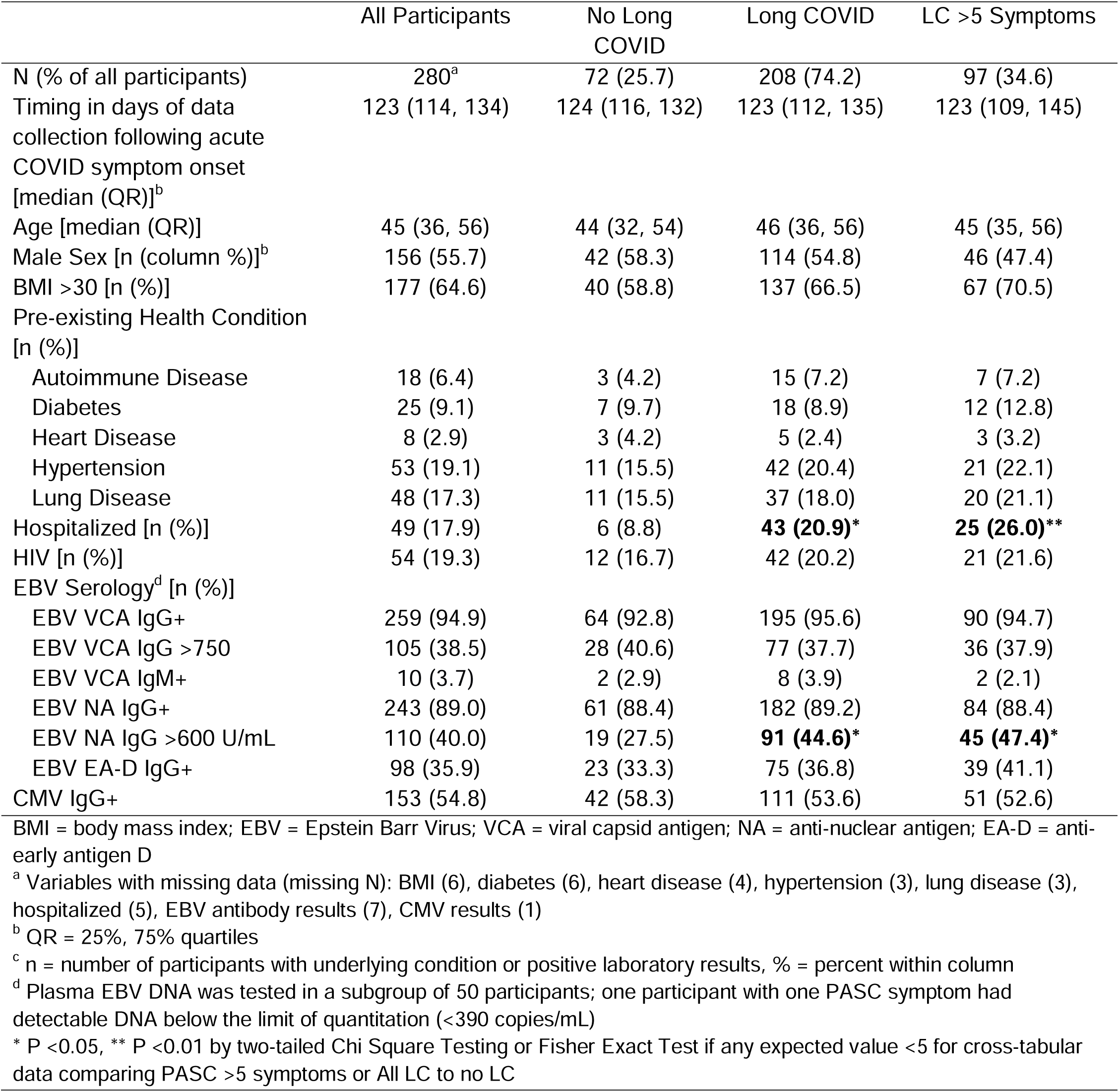
Participant demographics, pre-existing health conditions, prior hospitalization, and EBV serology results by Long COVID definitions including participant with sample time points greater than 60 days after initial infection.

|  | All Participants | No Long COVID | Long COVID | LC >5 Symptoms |
| --- | --- | --- | --- | --- |
| N (% of all participants) | 280<sup>a</sup> | 72 (25.7) | 208 (74.2) | 97 (34.6) |
| Timing in days of data collection following acute COVID symptom onset [median (QR)]<sup>b</sup> | 123 (114, 134) | 124 (116, 132) | 123 (112, 135) | 123 (109, 145) |
| Age [median (QR)] | 45 (36, 56) | 44 (32, 54) | 46 (36, 56) | 45 (35, 56) |
| Male Sex [n (column %)]<sup>b</sup> | 156 (55.7) | 42 (58.3) | 114 (54.8) | 46 (47.4) |
| BMI >30 [n (%)] | 177 (64.6) | 40 (58.8) | 137 (66.5) | 67 (70.5) |
| Pre-existing Health Condition [n (%)] |  |  |  |  |
| Autoimmune Disease | 18 (6.4) | 3 (4.2) | 15 (7.2) | 7 (7.2) |
| Diabetes | 25 (9.1) | 7 (9.7) | 18 (8.9) | 12 (12.8) |
| Heart Disease | 8 (2.9) | 3 (4.2) | 5 (2.4) | 3 (3.2) |
| Hypertension | 53 (19.1) | 11 (15.5) | 42 (20.4) | 21 (22.1) |
| Lung Disease | 48 (17.3) | 11 (15.5) | 37 (18.0) | 20 (21.1) |
| Hospitalized [n (%)] | 49 (17.9) | 6 (8.8) | **43 (20.9)\*** | **25 (26.0)\*\*** |
| HIV [n (%)] | 54 (19.3) | 12 (16.7) | 42 (20.2) | 21 (21.6) |
| EBV Serology<sup>d</sup> [n (%)] |  |  |  |  |
| EBV VCA IgG+ | 259 (94.9) | 64 (92.8) | 195 (95.6) | 90 (94.7) |
| EBV VCA IgG >750 | 105 (38.5) | 28 (40.6) | 77 (37.7) | 36 (37.9) |
| EBV VCA IgM+ | 10 (3.7) | 2 (2.9) | 8 (3.9) | 2 (2.1) |
| EBV NA IgG+ | 243 (89.0) | 61 (88.4) | 182 (89.2) | 84 (88.4) |
| EBV NA IgG >600 U/mL | 110 (40.0) | 19 (27.5) | **91 (44.6)\*** | **45 (47.4)\*** |
| EBV EA-D IgG+ | 98 (35.9) | 23 (33.3) | 75 (36.8) | 39 (41.1) |
| CMV IgG+ | 153 (54.8) | 42 (58.3) | 111 (53.6) | 51 (52.6) |

BMI = body mass index; EBV = Epstein Barr Virus; VCA = viral capsid antigen; NA = anti-nuclear antigen; EA-D = anti-early antigen D
<sup>a</sup> Variables with missing data (missing N): BMI (6), diabetes (6), heart disease (4), hypertension (3), lung disease (3), hospitalized (5), EBV antibody results (7), CMV results (1)
<sup>b</sup> QR = 25%, 75% quartiles
<sup>c</sup> n = number of participants with underlying condition or positive laboratory results, % = percent within column
<sup>d</sup> Plasma EBV DNA was tested in a subgroup of 50 participants; one participant with one PASC symptom had detectable DNA below the limit of quantitation (<390 copies/mL)
\* P <0.05, \*\* P <0.01 by two-tailed Chi Square Testing or Fisher Exact Test if any expected value <5 for cross-tabular data comparing PASC >5 symptoms or All LC to no LC

### Relationship between EBV serostatus *and Long COVID symptoms*

A higher proportion of participants who experienced LC or LC>5, compared with those without LC, had EBV NA IgG levels greater than the limit of quantitation of 600 U/mL (45% and 47% versus 28%; all P<0.05). While not significant in univariate analyses, we observed that participants with CMV seropositivity were less likely to have LC or LC with > 5 symptoms versus those without LC (54% and 53% versus 58%, respectively).

In order to determine the independent associations between demographic factors, pre-existing medical conditions and EBV NA and EA-D IgG results with LC and in those with specific LC symptoms, we performed covariate-adjusted binary logistic regression modeling as shown in **Figure 2** (adjusted for timing of sample collection >100 days, prior COVID-related hospitalization, age >50 years, sex, body mass index >30, pre-existing diabetes mellitus, hypertension, renal disease, and autoimmune disease, known HIV infection, CMV IgG seropositivity, EBV NA IgG >600 U/mL, and EBV EA-D IgG positivity). **Supplemental Figure 1** summarizes the number of symptoms experienced by each participant in the various LC symptom groups, which was roughly similar overall across symptom groups (median ranged from 8 to 9.5 with significantly higher number of symptoms in those with gastrointestinal symptoms compared with those with neurocognitive symptoms). 258 participants (92%) with data available across all variables were included in logistic regression. EBV antibody variables were selected for inclusion in the final regression models based on antibody measures that may represent recent EBV reactivation as recently reported (EBV EA-D IgG; (27)) or high levels of EBV NA IgG (i.e. >600 U/mL, the upper limit of assay detection) based on the association with LC in univariate analysis (**Table 1**). Notably, unlike detection of EA-D IgG from EBV reactivation, high levels of EBV NA IgG may either represent recent viral reactivation or be secondary to increased generalized inflammation from acute SARS-CoV-2 infection resulting in B cell activation and non-specific gammaglobulinemia. EBV NA IgG levels would be expected to peak months after reversion to the latent phase of EBV infection, around the time of sample collection in this study.

**Figure 2.**
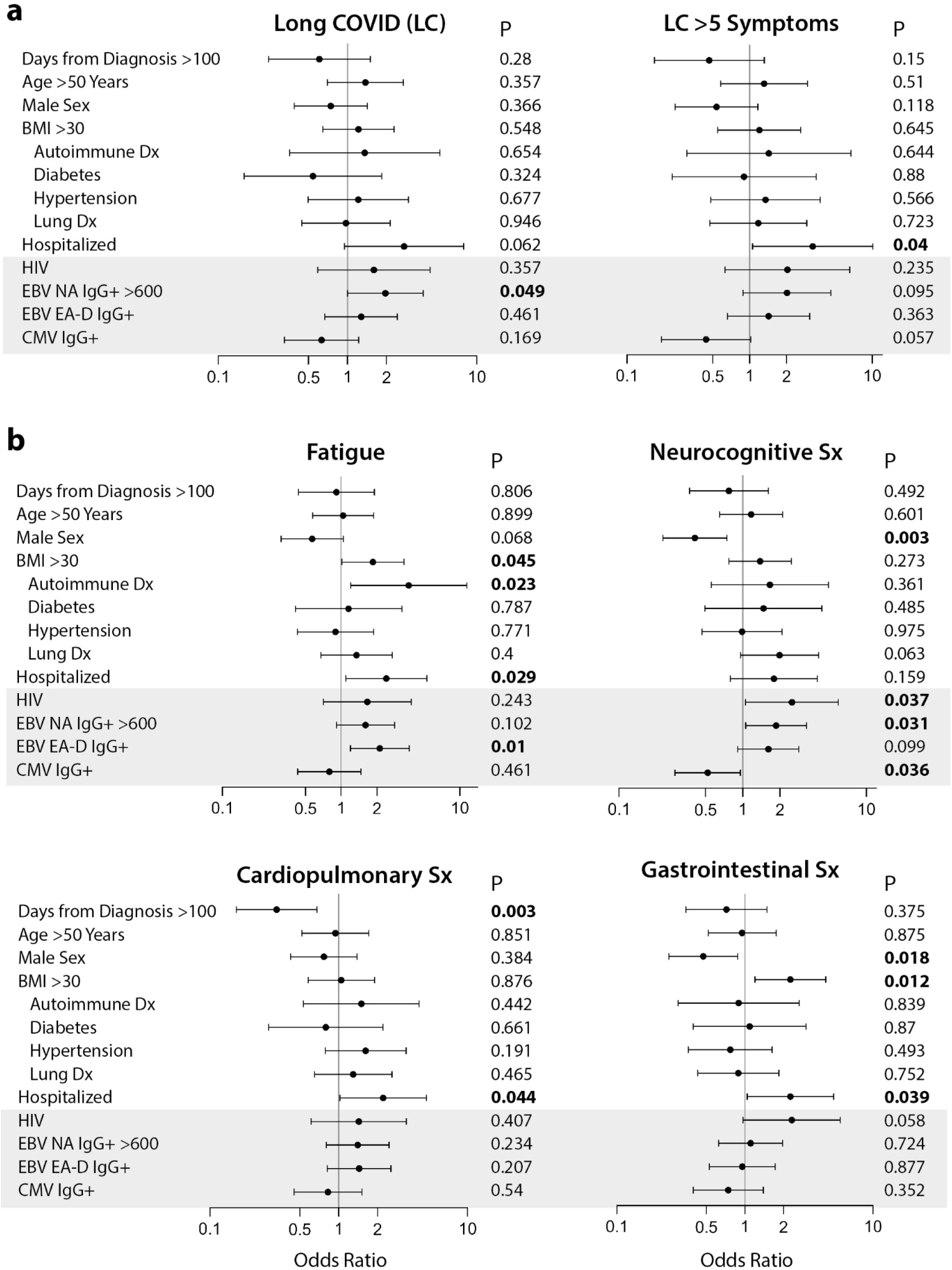
Results from covariate adjusted logistic regression analysis of predictors of Long COVID and long COVID symptoms. Demographic, underlying health conditions, HIV and CMV positivity, and EBV serological results as predictors of participants with any persistent symptom (PASC) or greater than 5 symptoms across organ systems compared with those without PASC are shown in (**a**). Associations between covariates and fatigue, neurocognitive symptoms (sx), cardiopulmonary symptoms, and gastrointestinal symptoms are shown in (**b**). N = 258 for all models with exception of LC >5 symptoms (N= 153; participants with one to four LC symptoms excluded from the analyses). Cases with missing values were excluded from the regression models. Dots and bars represent odds ratios and 95% confidence intervals. P values from regression analyses are shown adjusted for all covariates listed in the figure. BMI = body mass index; EBV = Epstein Barr Virus; VCA = viral capsid antigen; NA = nuclear antigen; EA-D = early antigen-D.; LC = long COVID.

EBV VCA IgG positivity, VCA IgG >limit of quantitation (750 U/mL), and VCA IgM results were not significant across any analyses and not included in the final models. Furthermore, very few participants had detectable VCA IgM levels (3.7%), which would be expected as sampling was conducted months after acute infection.

In adjusted regression analyses, the odds of LC>5, as well as LC characterized by fatigue, gastrointestinal symptoms, and cardiopulmonary symptoms were higher in those who had been hospitalized during acute infection (**Figure 2a-b**). Female sex also correlated with gastrointestinal and neurocongitive symptoms (**Figure 2b**).

Interestingly, participants reporting pre-existing autoimmune disease (mainly thyroiditis) and those who had detectable EBV EA-D IgG responses had a higher odds of experiencing fatigue (**Figure 2b**) a median of four months following COVID-19 diagnosis. Participants with high levels of EBV NA IgG levels (>600 U/mL) had higher odds of experiencing neurocognitive symptoms. Furthermore, the NA IgG >600 U/mL odds ratios were higher in those with any number of LC symptoms (**Figure 2a**); non-significant trends were observed for LC>5 symptoms and fatigue (**Figure 2a-b**).

### EBV DNA measurements

In order to determine if circulating EBV DNA is detectable during convalescence and whether any association between EBV DNA persistence and LC is present, we performed quantitative EBV PCR on plasma samples from a random subgroup of 50 participants who underwent EBV serological testing stratified by EA-D positivity (the subgroup demographics and participant phenotypes were similar to the larger cohort as shown in **Supplemental Table 1**). Only one of the fifty participants had detectable plasma EBV DNA, and the level was below the limit of quantitation (<390 copies/mL). This participant had no reported pre-existing medical conditions, had no detectable EA-D IgG or VCA IgM at the time of sampling, had EBV NA and VCA IgG greater than the limit of quantitation, and reported 2 LC symptoms (persistent cough and heart palpitations) at the time of sampling.

### Relationship between CMV serostatus and Long COVID symptoms

Next, we analyzed the impact of CMV seropositivity on LC symptom clusters in the same covariate-adjusted regression models as above for EBV (**Figure 2a-b**). CMV IgG positivity is not used to determine recent viral reactivation and is therefore solely a marker of pre-existing CMV infection. In contrast to EBV serological results, after adjustment for potential confounders, CMV seropositive participants had lower odds of developing neurocognitive LC (OR 0.52, P=0.036; **Figure 2b**) and exhibited trends towards lower odds of developing LC (OR 0.63, P=0.169) or LC>5 (OR 0.44, P=0.057), although these latter associations did not reach statistical significance (**Figure 2a**). There was no evidence for an association between CMV serostatus and fatigue or any of the other non-neurologic LC symptom clusters.

The lower odds of those with underlying CMV infection experiencing LC appeared to be out of proportion to the modestly lower percentages of those with LC who were CMV IgG positive (approximately 5% lower in those with LC or LC>5 than in participants without LC; **Table 1**). As a result, we repeated regression models with stepwise inclusion of covariates that may mask the negative association of CMV on LC symptoms. For example, the OR of developing neurocognitive PASC in those that were CMV seropositive was 0.87 (P=0.55) when only CMV was included in the model as the lone variable. With the addition of HIV, the OR decreased to 0.71 (P=0.21) and with addition of EBV EA-D IgG+ and EBV NA>600 U/mL the OR decreased to 0.75 (P=0.27). With HIV and EBV antibody results included, the OR further decreased to 0.63 (P=0.1). Addition of other variables had much more modest effects on the OR of CMV predicting neurocognitive LC, and with all covariates included in the model the OR was 0.52 (P=0.036) as in **Figure 2**.

### Relationship between HIV and Long COVID symptoms

Of participants with HIV, 8 had viral loads above the limit of quantification (40 copies/mL; quantifiable values were 41, 50, 78, 424, 750, 28118, 39267, 46745 copies/mL). The median CD4+ T cell count and percentage were 576 (404-785) and 32% (25-38%), respectively. The median CD4/CD8 ratio was 0.86 (0.51-1.2). Twelve individuals had CD4+ T cell counts below 350 cells/uL; of these, only 3 had CD4+ T cell counts below 200 cells/uL. Throughout all adjusted analyses, participants with pre-existing HIV tended to have higher odds of developing LC symptoms or LC symptom clusters, with a statistically significant association between HIV and neurocognitive LC (OR 2.5, P=0.037) and approaching significance with gastrointestinal symptoms (OR 2.33, P=0.058). We repeated covariate-adjusted analyses restricted to HIV-negative individuals (N=213) as all but one participant with HIV (98.2%) was CMV IgG positive, compared with 54.8% of total participants included in the LC analyses. EBV EA-D IgG positivity remained significantly and positively associated with fatigue (OR 2.26, P =0.012) and CMV seropositivity remained significantly and negatively associated with neurocognitive PASC (OR 0.51, P =0.037).

### Analyses of non-hospitalized participants

Many prior pathophysiological studies of post-acute sequelae have included a majority of participants who were hospitalized for acute COVID-19, with many receiving intensive care or mechanical ventilation. There may also be a survival bias of those who develop PASC after severe initial disease presentations. As a result, we next performed regression analyses restricted to participants that did not require hospitalization (N=211). Overall, the relationships observed in the total population between EBV and CMV serologies and other demographic factors and symptom clusters were similar. For example, the significant and positive association between EBV EA-D IgG and fatigue strengthened (OR 2.37, P =<0.001). The negative associations between CMV and neurocognitive symptoms (OR 0.53) and positive association between EBV NA>600 U/mL (OR 1.58) were similar to the entire cohort but lost statistical significance in the context of a smaller analysis population size.

### Association between EBV and CMV antibody results and circulating markers of inflammation

We previously identified significant correlations between various markers of inflammation and LC symptoms, such as IL-6 and TNFα (3, 33, 34). As a result, we examined the relationship between EBV and CMV antibody results in a subset of 143 participants (24 with HIV) who had circulating biomarker data as measured on the HD-X Simoa platform available including markers of neuronal injury, inflammation and immune activation (glial fibrillary acidic protein [GFAP, a marker of astrocyte activation], neurofilament light chain [NFL, a marker of neuronal injury], monocyte Chemoattractant Protein-1 [MCP-1], IFNγ, IL-6, IL-10, TNFα, and IP-10). We identified significantly higher levels of NFL and MCP-1 in participants with measurable EBV EA-D IgG and significantly higher levels of TNFα, IL-10 and MCP-1 in those with EBV VCA IgG >750 U/mL in two-sided, non-parametric analyses corrected for multiple comparisons (**Figure 3**). Despite having a substantially lower risk of neurocognitive LC, CMV seropositive participants had significantly higher plasma NFL, IL-6, IP-10, and TNFα levels than those without CMV (**Figure 3**).

**Figure 3.**
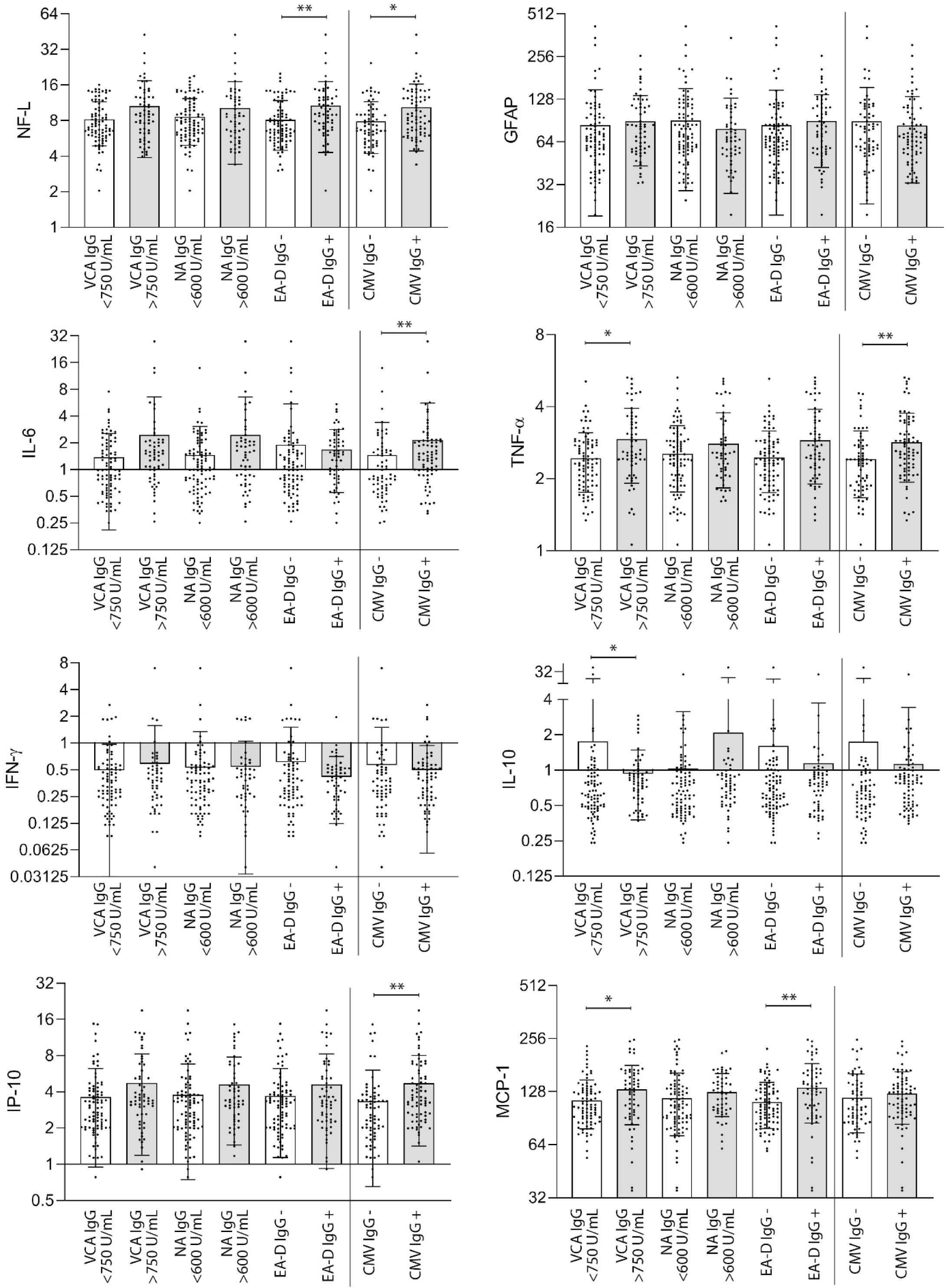
Circulating markers of Inflammation grouped by EBV and CMV antibody result. No significant differences in inflammation marker levels were observed within each antibody group (*e*.*g*. EA-D IgG + versus EA-D IgG -) by two-sided Kruskal-Wallis testing with Dunn’s correction for multiple comparison (* P <0.05, ** P <0.01). Bars and lines represent mean and standard deviation (all data points are shown). Units are in pg/mL.

### Impact of CMV on associations between circulating markers of inflammation and LC symptoms

Markers of inflammation, such as IL-6 and TNFα have been previously associated with LC/PASC and were elevated in participants with underlying CMV infection as above. However, CMV was negatively associated with LC outcomes in our regression modeling, and to help clarify the relationships between biomarkers and CMV as predictors of LC, we performed binary logistic regression including each biomarker alone or covariate adjusted with CMV IgG positivity with LC symptom clusters as shown in **Supplemental Table 2** (N=141 with all data available). Interestingly, adjusting for CMV status actually strengthened the associations between inflammation and Long COVID.

## DISCUSSION

In a cohort of several hundred individuals with confirmed prior SARS-CoV-2 infection, we found that certain factors associated with chronic viral infections, such as EBV reactivation and pre-existing HIV, were independently associated with various Long COVID symptom clusters. In contrast, participants who had serologic evidence of prior CMV infection were less likely to report neurocognitive symptoms and tended to have less LC overall. Furthermore, HIV, EBV EA-D IgG positivity and high titers of EBV NA IgG appeared to mask the negative effects of CMV on LC. Of note, we identified LC even those without evidence of EBV reactivation or CMV disease, suggesting that these factors are not essential to the development of persistent symptoms or sequelae.

Our study confirms and extends prior studies that identified an association between EBV EA-D positivity and LC symptoms, raising the intriguing hypothesis that EBV reactivation may be mechanistically related to specific LC syndromic phenotypes. By carefully defining LC syndromic phenotypes and adjusting for various participant factors, sample timing, underlying health conditions and prior hospitalization, we identified a strong association between evidence of recent EBV reactivation and fatigue, one of the most prevalent LC symptoms. We were able to demonstrate that serologic EBV reactivation may be specifically associated with fatigue and neurologic symptoms, but less so with other LC syndromic phenotypes (*i*.*e*., cardiopulmonary, gastrointestinal). In analyses excluding participants that were hospitalized, we were able to confirm that these associations are not entirely due to differences in acute COVID-19 severity. Whether or not EBV reactivation is the root cause of these symptoms, it should be noted that primary EBV infection (*e*.*g*., mononucleosis) may lead to prolonged fatigue, and EBV seroconversion has recently been shown to be common prior to the development of MS, an autoimmune condition that may be precipitated by aberrant, autoreactive immune responses to this virus (20). Since autoimmunity has been proposed as pathophysiologic mechanisms underlying LC (7, 15) and pre-existing autoimmunity was associated with LC in our analysis, further study of its potential relationship with EBV disease activity in this patient population is warranted.

The biological mechanisms leading to high levels of EBV NA IgG (greater than the assay limit of detection of 600 U/mL) observed in association with LC symptoms and neurocognitive symptoms is not entirely clear. Whereas EA-D IgG responses are generally understood to be a result of recent EBV reactivation in those with pre-existing latent EBV infection (27), nearly 90% of our cohort had detectable NA IgG, consistent with the long-lasting nature of this antibody and high proportion of participants with pre-existing EBV infection. It is possible that those with higher levels experienced a recent increase following EBV reactivation, but given the lack of sampling during or before acute SARS-CoV-2 infection, we do not know for certain. Nonetheless, NA IgG responses usually peak during establishment (or perhaps re-establishment) of EBV latency (16, 17, 28), the timing of which is consistent with the post-acute sample collection timing here. It is also possible that high EBV NA IgG levels resulted from non-specific hypergammaglobulinemia that can develop during acute viral infections. Further studies in convalescent cohorts with samples collected during acute infection are urgently needed.

We made the surprising and novel observation that CMV seropositivity was negatively associated with the development of Long COVID phenotypes. The mechanism underlying this observation is not immediately clear, and we can only speculate on possible explanations. It is plausible that CMV seropositive individuals might mount more robust adaptive immune responses to SARS-CoV-2. For example, CMV seropositivity in younger adults is actually associated with heightened adaptive immune responses to influenza vaccination (35), despite earlier studies in the aging literature linking CMV to immunosenescence phenotypes (36). Alternatively, CMV-induced immunoregulatory pathways, including secretion of its own viral IL-10, might dampen local inflammation in areas of CMV reactivation, decreasing the risk of auto-antibody formation (to the extent that autoantibodies may contribute to the risk of neurologic LC symptoms) (37, 38). It is also unclear whether these associations reflect a direct causal effect of CMV on LC risk or host factors that affect the risk of CMV infection and LC independently. It is interesting that CMV serostatus was more strongly associated with neurologic LC symptoms than other syndromic phenotypes. While CMV-infected myeloid cells can be found in the central nervous system and CMV-induced inflammation might plausibly affect blood-brain barrier permeability (39), it is not immediately clear why CMV status would be so specifically linked to neurologic as opposed to non-neurologic LC symptoms. Lastly, why two chronic herpesvirus infections - EBV and CMV - have qualitatively different associations with LC remains entirely unclear, though perhaps the anatomic localization of herpesvirus reactivation is an important factor. For example, EBV preferentially reactivates within B cell follicles, where antibody responses develop, while CMV preferentially reactivates elsewhere (40).

It is particularly interesting that CMV seropositivity is associated with decreased odds of developing LC risk but worse disease severity in acute COVID-19, as reported in some recent studies (29, 30). Although CMV seropositivity was not completely protective against Long COVID in our study, the differential effects of CMV serostatus on acute versus Long COVID suggests that assessment of CMV serostatus may be important in future mechanistic evaluations of COVID-19. Indeed, since CMV seropositivity is associated with increased systemic inflammation, but a decreased risk of Long COVID, adjusting for CMV serostatus actually strengthened our previously reported associations between systemic inflammation and Long COVID symptoms (3, 33). This finding suggests that sources of inflammation unrelated to CMV are most likely driving PASC risk in COVID-19 survivors and highlights the importance of the source of inflammation - as opposed to simply systemic inflammation itself - in mediating the risk of PASC.

It is also notable that HIV was independently associated with the development of neurologic LC, and to a lesser degree gastrointestinal symptoms, than other LC syndromic phenotypes (*e*.*g*., fatigue, which was more closely linked to EBV reactivation). Thus, each chronic viral infection assessed in our study not only affected the risk of LC, but also exhibited specific and distinct syndromic associations. Whichever mechanisms explain these findings, these observations highlight the importance of measuring specific LC syndromic phenotypes as their underlying pathogenic mechanisms may well be distinct. They also highlight the likely heterogeneous nature of LC and may help determine inclusion in various future interventional trials. In fact, it will likely be difficult to prove any causal or modifying role of LC (*e*.*g*., EBV reactivation, CMV serostatus, long-term SARS-CoV-2 viral persistence, autoreactive immunity, etc.) without measuring the effects of targeted interventions in well-designed studies. Furthemore, given that there is paucity of circulating EBV during convalescence, the potential impact of EBV reactivation on the development of LC is likely to be greatest during acute COVID-19, and factors such as this will need to be considered in the design of such interventional studies.

Strengths of this study include the large sample of well-characterized post-acute COVID-19 patients, most of whom were not hospitalized during acute infection, at a time point consistent with consensus case definitions of Long COVID. Nevertheless, the study has several limitations. Although diverse, our cohort is a convenience sample not representative of all individuals with COVID-19 or Long COVID. In particular, while we specifically oversampled people with treated HIV infection to assess its association with Long COVID, we have a limited subsample of people with HIV to detect modest effect sizes. We also did not have access to biospecimens from acute or very early convalescent infection (<30 days). Direct evaluation of EBV dynamics during these early phases is warranted, although we believe our results strongly suggest that investigation of EBV viremia during post-acute stages is of limited utility. Finally, EBV and CMV reactivation are often tissue-based processes and such samples may be needed in order to identify persistent, smoldering infection. As a result, tissue studies will be critical to understanding the full pathophysiological mechanisms underlying LC.

In summary, this study expands our understanding of the relationships between chronic viral infections and the risk of distinct LC syndromic phenotypes. While it remains unclear whether these associations reflect causal effects of viral co-infections or host factors associated with viral co-infections on LC, these observations suggest distinct pathogenesis of the various LC syndromic phenotypes. We also extend prior reports that serological evidence of recent EBV reactivation is associated with LC, by demonstrating that these associations primarily involve fatigue and neurologic LC symptoms. We also made the novel observation that CMV seropositivity has an unexpected, negative association with LC, which in turn, is masked to some degree by HIV infection and EBV reactivation. Nevertheless, the presence of LC symptoms could not be completely explained by the viral co-infections assessed in our study, suggesting that other factors must be important mediators of LC. In particular, it remains to be seen whether SARS-CoV-2 persistence in tissues may also play a role in LC as suggested by recent uncontrolled case series of SARS-CoV-2-directed antiviral therapies (41–43). Ultimately, further investigation of SARS-CoV-2 and other viruses during both acute infection and convalescence will be needed to clarify the mechanisms driving Long COVID and suggest interventions that may reverse or ameliorate these processes.

## MATERIALS & METHODS

### Study participants

All participants in the Long-term Impact of Infection with Novel Coronavirus cohort (LIINC; NCT04362150) with biospecimens available outside the acute window of SARS-CoV-2 infection were studied; the cohort procedures have been described in detail previously (44). Briefly, any adult with a history of SARS-CoV-2 infection identified on nucleic acid amplification testing, regardless of the presence of acute or post-acute symptoms, was eligible to enroll >14 days following symptom onset and followed approximately every 4 months thereafter. Participants were recruited through a combination of mailings to all individuals testing positive at two academic medical centers as well as clinician- and self-referrals, as described elsewhere (44). We also deliberately enriched the cohort for people with HIV by notifying all eligible individuals testing positive for COVID-19 at two university-affiliated HIV clinics to allow us to assess the association between HIV and LC symptoms.

Data regarding the acute period of COVID-19 (including number, type, and severity of symptoms, hospitalization and COVID-19 treatment), as well as demographics, and medical comorbidities, were collected by self-report at the first visit and verified through review of medical records whenever possible. At each visit, participants were queried regarding the presence of 32 symptoms derived from the U.S. Centers for Disease Control COVID-19 symptom list (45) and the Patient Health Questionnaire (PHQ) somatic symptom scale (46). Importantly, participants were specifically asked to describe symptoms only if they were new or worse compared to the period prior to COVID-19 (pre-existing symptoms were not considered to represent LC). Participants were also asked to assign themselves a score using a visual-analogue scale from 0-100 to indicate their overall health prior to COVID-19, at the worst point in their illness, and in the week prior to the visit.

### Biospecimen collection

At each visit, whole blood was collected in EDTA tubes followed by density gradient separation and isolation of peripheral blood mononuclear cells and plasma as previously described (47). Serum was obtained concomitantly from serum-separation tubes for antibody testing. Both plasma and serum samples were stored at -80F.

### EBV assays

EBV antibody testing was performed on participant serum by ARUP laboratories. The EBV antibody panel included quantitative measures of anti-Viral Capsid Antigen (VCA) IgG and IgM, anti-Nuclear Antigen (NA) IgG, and early antigen-diffuse IgG. Results were considered positive in this analysis if units (U) per mL were within or higher than the indeterminate range of the assay (VCA IgG ≥ 18 U/mL; VCA IgM ≥ 36 U/mL; NA IgG ≥ 18 U/mL; early D Ag ≥ 9 U/mL). The VCA IgG, NA IgG and EA-D IgG assays had upper limits of quantitation (>750 U/mL, >600 U/mL and >150 U/mL, respectively). Quantitative EBV PCR testing was performed on a random subset of 50 study participants stratified by EA-D IgG positivity by ARUP laboratories (quantitative range 2.6-7.6 log copies/mL). This assay also identifies detectable EBV DNA above and below the limit of quantitation.

### CMV assays

CMV serostatus was assessed in duplicate on cryopreserved serum by qualitative ELISA (CMV IgG ELISA [GWB-BQK12C], Genway Biotech, San Diego, CA), with antibody index values <0.9 considered negative, >1.1 considered positive, and between 0.9 and 1.1 considered indeterminate per manufacturer specifications. Levels greater than 0.9 were considered detectable in this study. For participants without available serum at study entry, subsequent visits up to 20 months following COVID-19 diagnosis were used for serostatus ascertainment as while the prevalence of CMV is high in the general population, the incidence among seronegative adults is typically <1% per year (48).

### Biomarker and SARS-CoV-2 IgG analyses

A subset of participants (n=143) had circulating biomarker data available from previous testing (3, 49). Briefly, the fully automated HD-X Simoa platform was used to measure biomarkers in blood plasma including monocyte chemoattractant protein-1 (MCP-1), Cytokine 3-PlexA (interleukin 6 [IL-6], interleukin 10 [IL-10], tumor necrosis factor alpha [TNF-α]), IFN-γ–induced protein 10 (IP-10), IFN-γ, neurofilament light chain (NfL), glial fibrillary acidic protein (GFAP), and SARS-CoV-2 receptor-binding domain (RBD) immunoglobulin G (IgG) according to the manufacturer’s instructions. Assay performance was consistent with the manufacturer’s specifications.

### Statistical methods

Descriptive statistics were used to characterize the cohort including median and 25% and 75% quartiles for continuous variables. In univariate analyses of binary variables, we performed two-sided chi square testing or Fisher’s exact testing (if any expected cell value was less than 5) for cross-tabular data and two-sided Mann-Whitney U or Kruskal-Wallis tests (for multiple comparisons with Dunn correction) to compare variables across Long COVID groups, symptom groups, and EBV antibody results. Covariate-adjusted binary logistic regression models were performed to determine independent associations between variables and PASC/symptom/antibody results. Continuous biomarker data used in binary regression models were log_10_ transformed to achieve normality and divided by the IQR for each individual biomarker in order normalize the effect size across variables. All P values are 2 sided. Prism version 9.1.2 (GraphPad Software, San Diego, California) and SPSS version 28.0.1.1 (IBM) software was used for analyses.

### Human subjects

All participants provided written informed consent. The study was approved by the Institutional Review Board at the University of California, San Francisco.

## Supporting information

LIINC EBV CMV Supplement

## Data Availability

All data produced in the present study are available upon reasonable request to the authors.

## FOOTNOTES

## Acknowledgements

We are grateful to the LIINC study participants and to the clinical staff who provided care to these individuals during their acute illness period and during their recovery. We thank Dr. Isabel Rodriguez-Barraquer, Dr. Bryan Greenhouse, and Dr. Rachel Rutishauser for their contributions to the LIINC leadership team. We thank Elnaz Eilkhani for coordination with the Institutional Review Board. We acknowledge the contributions of the UCSF Clinical and Translational Science Unit, Core Immunology Laboratory, and AIDS Specimen Bank.

## Author Contributions

MJP, SGD, PWH, and TJH designed the study, which was supported through funding to MAS, JDK, JNM, SGD, PWH, and TJH. MJP, RH, JDK, JNM, SGD, and TJH designed the cohort. DR, SEM, RH, VT, LT, MD, LHN, MB, and AR collected clinical data and biospecimens, which were processed by TMD, DR, SEM, LT, NI in the laboratory of TJH. Specimens were analyzed by TMD, DR, SEM, GB-E, FC, BCY, AC, JWW, and CJP. SL, SAG, JYC processed and managed data. MJP and TJH performed and/or interpreted the statistical analyses. MJP, SGD, PWH, and TJH drafted the initial manuscript with input from AD, JH, MAS, MSD, PYH, JDK, and JNM. All authors edited, reviewed, and approved the final manuscript.

---

**Supplemental Figure 1.**
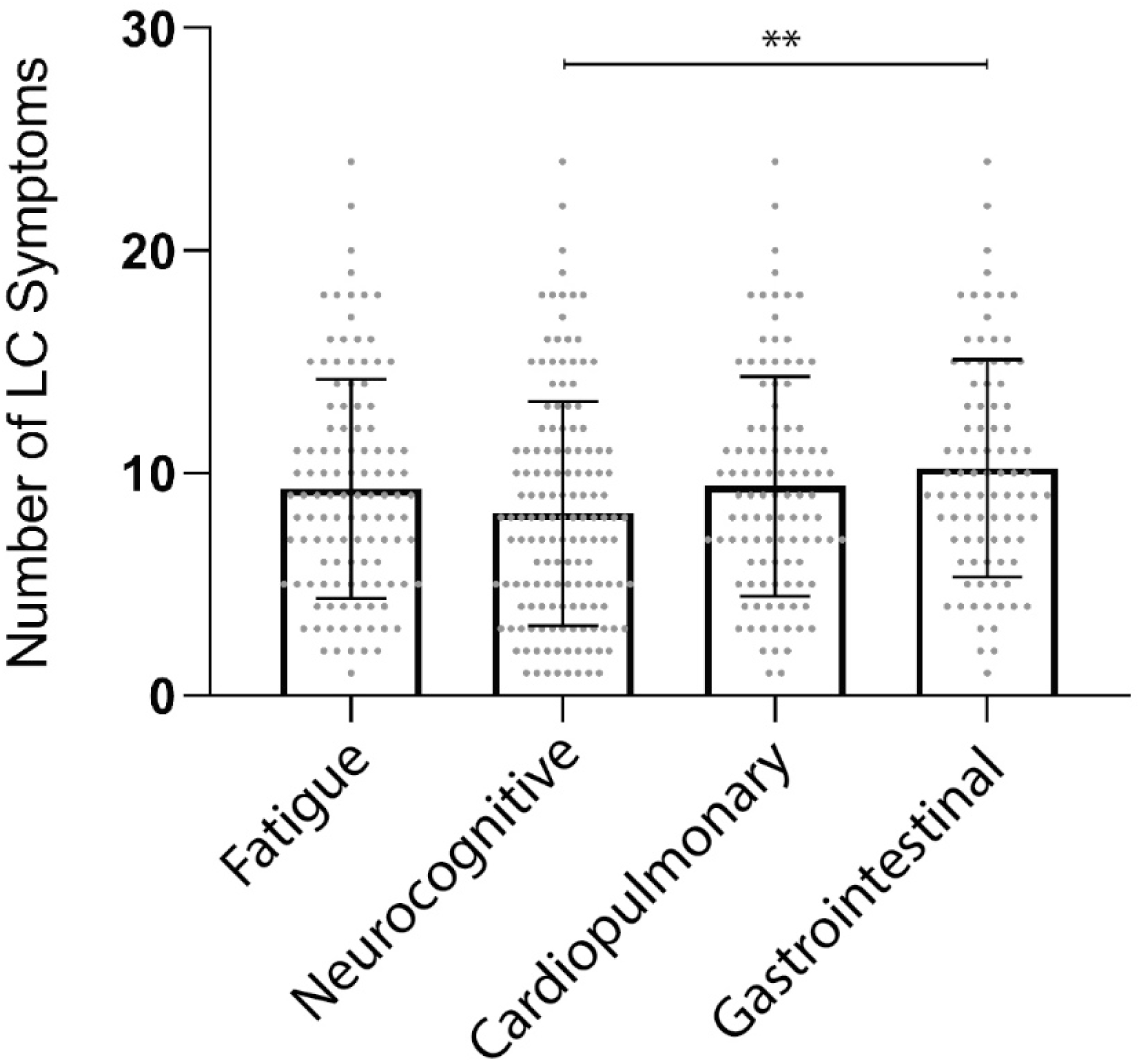
Number of Long COVD (LC) symptoms at the time of sample collection (median 4 months following SARS-CoV-2 PCR diagnosis) by LC symptom phenotype. Bars and lines represent mean and standard deviation. ** P <0.01 as determine by Kruskal-Wallis test with Dunn correction for multiple comparisons. Each point represents a study participant.

**Supplemental Table 1.**
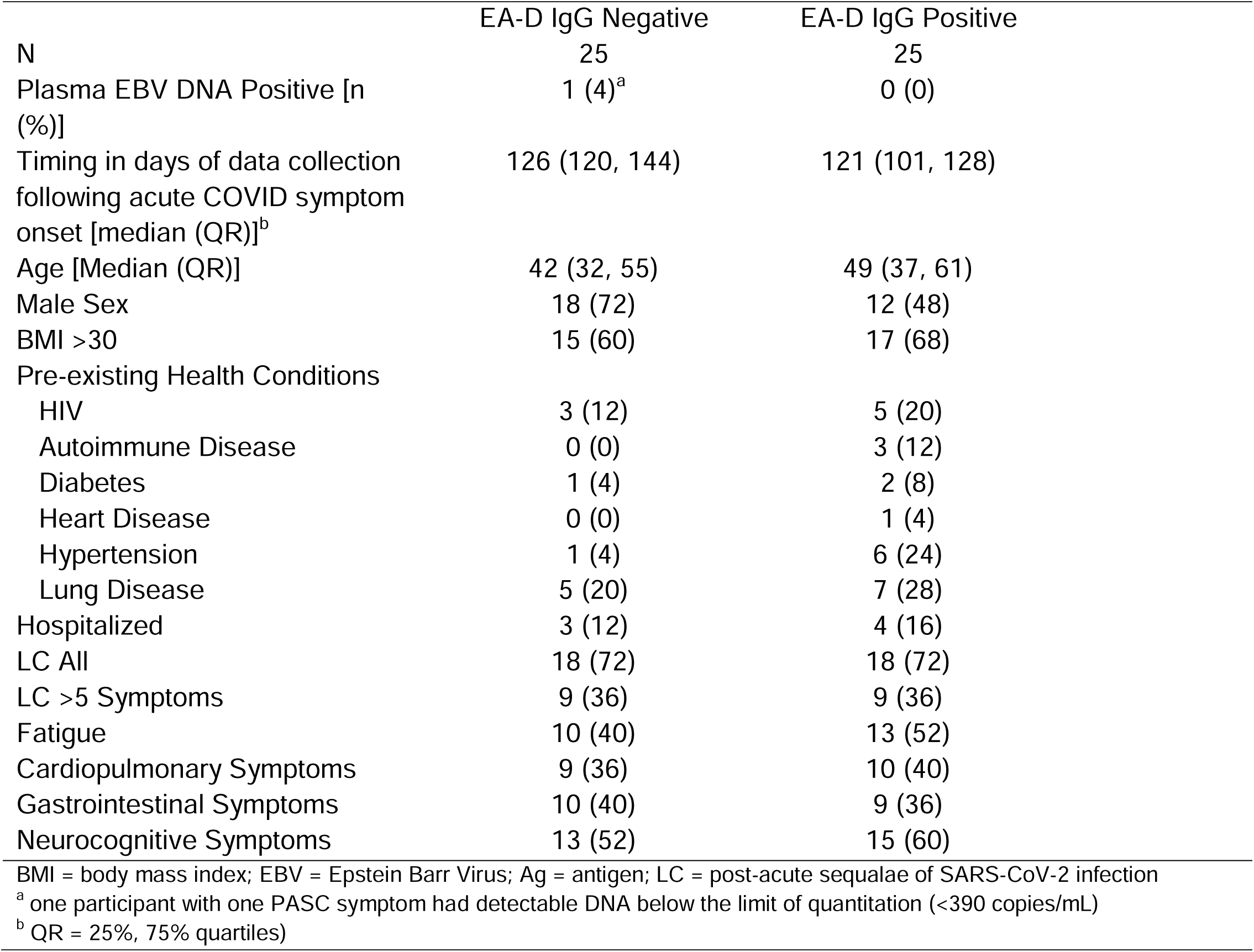
Subgroup analysis of plasma EBV DNA testing and other factors in subgroups of randomly selected participants stratified by Early Antigen-Diffuse (EA-D) IgG positivity.

**Supplemental Table 2.**
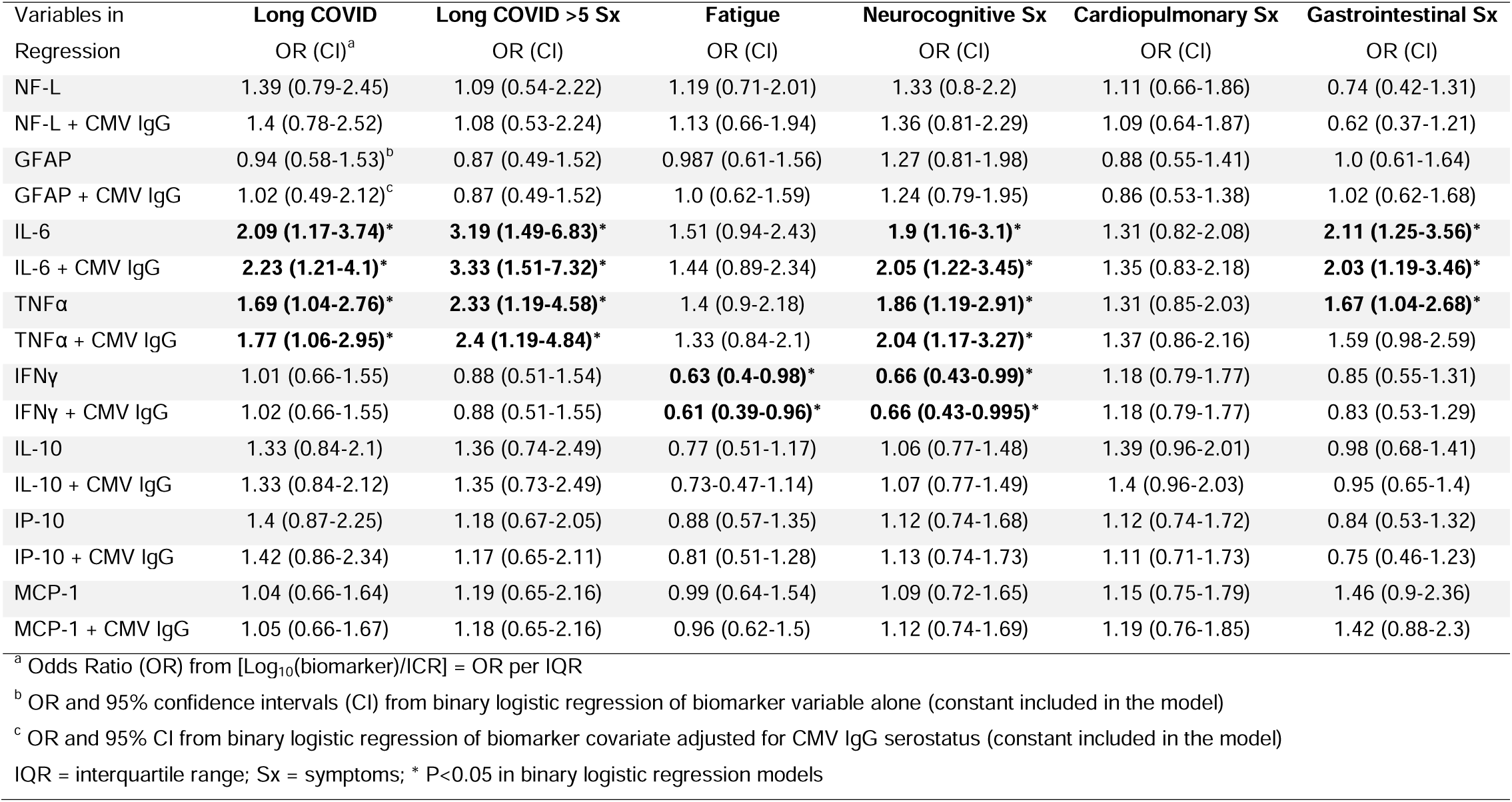
Binary logistic regression results of circulating markers of inflammation by Long COVID symptom clusters with and without adjusting for CMV IgG results.

