## Supplementary material for "Impact of Pre-Existing Chronic Viral Infection and Reactivation on the Development of Long COVID": LIINC EBV CMV Supplement

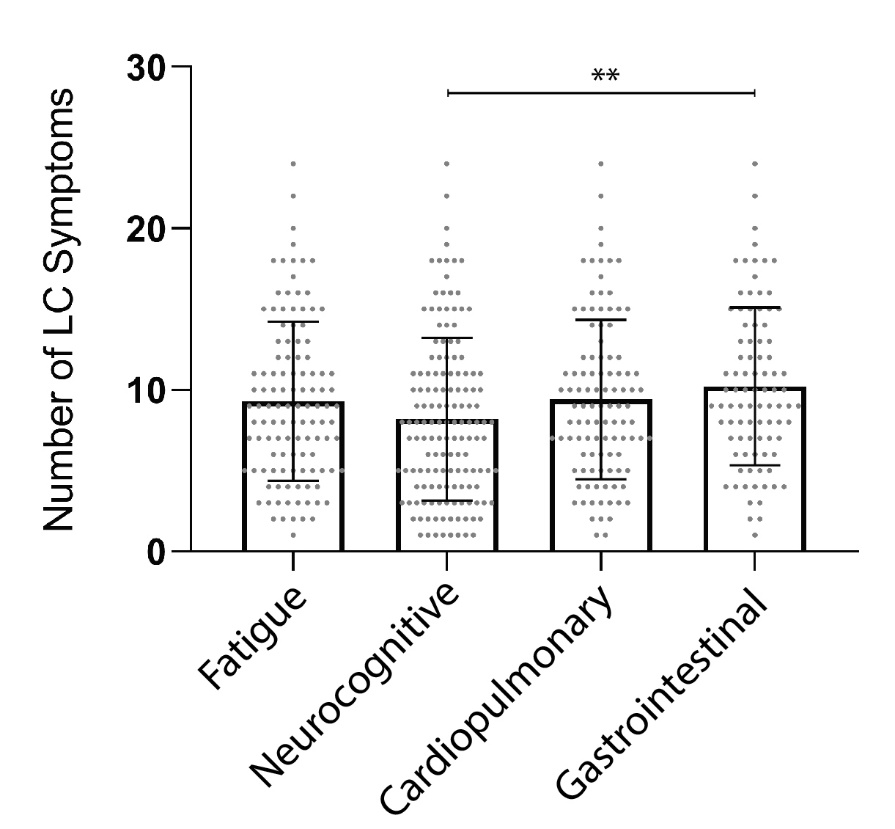

**Supplemental Figure 1**. Number of Long COVD (LC) symptoms at the time of sample collection (median 4 months following SARS-CoV-2 PCR diagnosis) by LC symptom phenotype. Bars and lines represent mean and standard deviation. ** P <0.01 as determine by Kruskal-Wallis test with Dunn correction for multiple comparisons. Each point represents a study participant.

| **Supplemental Table 1**. Subgroup analysis of plasma EBV DNA testing and other factors in subgroups of randomly selected participants stratified by Early Antigen-Diffuse (EA-D) IgG positivity. | | |
| --- | --- | --- |
|  | EA-D IgG Negative | EA-D IgG Positive |
| N | 25 | 25 |
| Plasma EBV DNA Positive [n (%)] | 1 (4)^a^ | 0 (0) |
| Timing in days of data collection following acute COVID symptom onset [median (QR)]^b^ | 126 (120, 144) | 121 (101, 128) |
| Age [Median (QR)] | 42 (32, 55) | 49 (37, 61) |
| Male Sex | 18 (72) | 12 (48) |
| BMI >30 | 15 (60) | 17 (68) |
| Pre-existing Health Conditions |  |  |
| HIV | 3 (12) | 5 (20) |
| Autoimmune Disease | 0 (0) | 3 (12) |
| Diabetes | 1 (4) | 2 (8) |
| Heart Disease | 0 (0) | 1 (4) |
| Hypertension | 1 (4) | 6 (24) |
| Lung Disease | 5 (20) | 7 (28) |
| Hospitalized | 3 (12) | 4 (16) |
| LC All | 18 (72) | 18 (72) |
| LC >5 Symptoms | 9 (36) | 9 (36) |
| Fatigue | 10 (40) | 13 (52) |
| Cardiopulmonary Symptoms | 9 (36) | 10 (40) |
| Gastrointestinal Symptoms | 10 (40) | 9 (36) |
| Neurocognitive Symptoms | 13 (52) | 15 (60) |
| BMI = body mass index; EBV = Epstein Barr Virus; Ag = antigen; LC = post-acute sequalae of SARS-CoV-2 infection  ^a^ one participant with one PASC symptom had detectable DNA below the limit of quantitation (<390 copies/mL)  ^b^ QR = 25%, 75% quartiles) | | |

| **Supplemental Table 2.** Binary logistic regression results of circulating markers of inflammation by Long COVID symptom clusters with and without adjusting for CMV IgG results. | | | | | | |
| --- | --- | --- | --- | --- | --- | --- |
| Variables in Regression | **Long COVID** OR (CI)^a^ | **Long COVID >5 Sx** OR (CI) | **Fatigue** OR (CI) | **Neurocognitive Sx** OR (CI) | **Cardiopulmonary Sx** OR (CI) | **Gastrointestinal Sx** OR (CI) |
| NF-L | 1.39 (0.79-2.45) | 1.09 (0.54-2.22) | 1.19 (0.71-2.01) | 1.33 (0.8-2.2) | 1.11 (0.66-1.86) | 0.74 (0.42-1.31) |
| NF-L + CMV IgG | 1.4 (0.78-2.52) | 1.08 (0.53-2.24) | 1.13 (0.66-1.94) | 1.36 (0.81-2.29) | 1.09 (0.64-1.87) | 0.62 (0.37-1.21) |
| GFAP | 0.94 (0.58-1.53)^b^ | 0.87 (0.49-1.52) | 0.987 (0.61-1.56) | 1.27 (0.81-1.98) | 0.88 (0.55-1.41) | 1.0 (0.61-1.64) |
| GFAP + CMV IgG | 1.02 (0.49-2.12)^c^ | 0.87 (0.49-1.52) | 1.0 (0.62-1.59) | 1.24 (0.79-1.95) | 0.86 (0.53-1.38) | 1.02 (0.62-1.68) |
| IL-6 | **2.09 (1.17-3.74)*** | **3.19 (1.49-6.83)*** | 1.51 (0.94-2.43) | **1.9 (1.16-3.1)*** | 1.31 (0.82-2.08) | **2.11 (1.25-3.56)*** |
| IL-6 + CMV IgG | **2.23 (1.21-4.1)*** | **3.33 (1.51-7.32)*** | 1.44 (0.89-2.34) | **2.05 (1.22-3.45)*** | 1.35 (0.83-2.18) | **2.03 (1.19-3.46)*** |
| TNFα | **1.69 (1.04-2.76)*** | **2.33 (1.19-4.58)*** | 1.4 (0.9-2.18) | **1.86 (1.19-2.91)*** | 1.31 (0.85-2.03) | **1.67 (1.04-2.68)*** |
| TNFα + CMV IgG | **1.77 (1.06-2.95)*** | **2.4 (1.19-4.84)*** | 1.33 (0.84-2.1) | **2.04 (1.17-3.27)*** | 1.37 (0.86-2.16) | 1.59 (0.98-2.59) |
| IFNγ | 1.01 (0.66-1.55) | 0.88 (0.51-1.54) | **0.63 (0.4-0.98)*** | **0.66 (0.43-0.99)*** | 1.18 (0.79-1.77) | 0.85 (0.55-1.31) |
| IFNγ + CMV IgG | 1.02 (0.66-1.55) | 0.88 (0.51-1.55) | **0.61 (0.39-0.96)*** | **0.66 (0.43-0.995)*** | 1.18 (0.79-1.77) | 0.83 (0.53-1.29) |
| IL-10 | 1.33 (0.84-2.1) | 1.36 (0.74-2.49) | 0.77 (0.51-1.17) | 1.06 (0.77-1.48) | 1.39 (0.96-2.01) | 0.98 (0.68-1.41) |
| IL-10 + CMV IgG | 1.33 (0.84-2.12) | 1.35 (0.73-2.49) | 0.73-0.47-1.14) | 1.07 (0.77-1.49) | 1.4 (0.96-2.03) | 0.95 (0.65-1.4) |
| IP-10 | 1.4 (0.87-2.25) | 1.18 (0.67-2.05) | 0.88 (0.57-1.35) | 1.12 (0.74-1.68) | 1.12 (0.74-1.72) | 0.84 (0.53-1.32) |
| IP-10 + CMV IgG | 1.42 (0.86-2.34) | 1.17 (0.65-2.11) | 0.81 (0.51-1.28) | 1.13 (0.74-1.73) | 1.11 (0.71-1.73) | 0.75 (0.46-1.23) |
| MCP-1 | 1.04 (0.66-1.64) | 1.19 (0.65-2.16) | 0.99 (0.64-1.54) | 1.09 (0.72-1.65) | 1.15 (0.75-1.79) | 1.46 (0.9-2.36) |
| MCP-1 + CMV IgG | 1.05 (0.66-1.67) | 1.18 (0.65-2.16) | 0.96 (0.62-1.5) | 1.12 (0.74-1.69) | 1.19 (0.76-1.85) | 1.42 (0.88-2.3) |
| ^a^ Odds Ratio (OR) from [Log_10_(biomarker)/ICR] = OR per IQR  ^b^ OR and 95% confidence intervals (CI) from binary logistic regression of biomarker variable alone (constant included in the model)  ^c^ OR and 95% CI from binary logistic regression of biomarker covariate adjusted for CMV IgG serostatus (constant included in the model)  IQR = interquartile range; Sx = symptoms; * P<0.05 in binary logistic regression models | | | | | | |
